# Social Determinants of Health Associated with Multiple Myeloma Incidence and Survival among a Low-Income Cohort in the Southeastern U.S

**DOI:** 10.64898/2026.01.02.26343344

**Authors:** Anna Junkins, Wanqing Wen, Loren Lipworth, Xijing Han, Heather Munro, Michael T. Mumma, Martha J. Shrubsole, Wei Zheng, Eden Biltibo, Staci Sudenga

**Affiliations:** Division of Epidemiology, Department of Medicine, Vanderbilt University Medical Center, Nashville, TN, USA; Vanderbilt-Ingram Cancer Center, Nashville, TN, USA; Division of Hematology, Department of Medicine, Vanderbilt University Medical Center, Nashville, TN, USA

**Author notes:** **Corresponding Author**: Staci L. Sudenga, Ph.D., Vanderbilt University Medical Center 2525 West End Ave, Suite 800, Nashville, TN 37203.

**Keywords:** social determinants of health, multiple myeloma, residential racial segregation

## Abstract

**Objectives:** Multiple myeloma (MM) is the second most common hematologic malignancy in the U.S.; however, the etiology is poorly understood. We investigated social determinants of health (SDoH) associated with MM incidence and survival among low-income Black and White participants in the Southern Community Cohort Study (SCCS).

**Methods:** The SCCS enrolled participants aged 40-79 years from 12 Southeastern states. We examined associations between SDoH (residential racial segregation, neighborhood deprivation, population density, persistent poverty, and rurality) geocoded to zip code, with MM incidence and all-cause mortality using multivariable Cox regression analyses. Additional stratified analyses examined MM incidence by obesity status.

**Results:** Among 74,294 participants, there were 162 MM cases, 133 self-identified as Black individuals and 29 self-identified as White individuals. Living in the highest vs. lowest deprivation areas was associated with 2-fold increased MM risk (HR:2.24; 95% CI:1.01-4.95). Among MM cases, those living in the least vs. most residentially segregated areas had 2-fold increased mortality (HR:2.21; 95% CI:1.03-4.74). Among participants who were not obese, those who lived in the most densely populated areas had a reduced risk of MM compared to those who lived in the least densely populated areas (HR:0.31; 95% CI: 0.14-0.67); and those who lived in urban vs. rural areas had a reduced risk (HR:0.52; 95% CI: 0.32-0.85).

**Discussion:** SDoH factors including neighborhood deprivation and residential racial segregation could influence MM risk and survival among low-income populations.

**Conclusion:** SDOH factors should be considered when developing strategies to reduce overall MM burden, and disparities among people with lower socioeconomic backgrounds.

## 1. Introduction

Multiple myeloma (MM), a cancer of the plasma cells, is the second most common hematologic cancer in the U.S., with an estimated 36,110 new cases to be diagnosed in 2025. [1] Accounting for 2% of cancer-related mortality in the U.S., there will be an estimated 12,030 deaths caused by MM in 2025. [1] The relative 5-year survival for individuals with MM in the U.S. is about 62%. Incidence rates and mortality rates among non-Hispanic Black individuals are over 2-fold higher compared to the rates among their non-Hispanic White counterparts, and the majority of MM cases are diagnosed among people ≥65 years of age. [1] There are several established risk factors, including obesity and excess weight, family history of MM, and the presence of other plasma cell diseases, including the precursor monoclonal gammopathy of undetermined significance (MGUS), and solitary plasmacytoma; however, the etiology of MM is poorly understood. [2]

Social factors may influence MM risk, and unequal access to care can worsen MM outcomes. [3–6] Social determinants of health (SDoH), defined by the World Health Organization as “conditions in which people are born, grow, work, live and age, and the wider set of forces and systems shaping the conditions of daily life,” include factors such as education, income, insurance coverage, neighborhood deprivation. [7, 8] Having lower income and living in areas with higher deprivation have been associated with worse clinical outcomes among people with MM. [6] Furthermore, residential racial segregation is associated with racial disparities observed across many health conditions. [9–11] However, the impact of SDoH on MM outcomes among people living in the U.S. South, which experiences substantial poverty, has not been comprehensively investigated. [12] The objective of the current study is to fill this research gap by identifying SDoH factors associated with MM incidence and survival among a predominately low-income, urban-dwelling population in the Southeastern U.S.

## 2. Materials and Methods

### 2.1. Participant population and data source

We retrospectively analyzed data from the Southern Community Cohort Study (SCCS), a large, prospective cohort study, which includes about 85,000 (>60% Non-Hispanic Black/African American) adults, living in 12 states in the Southeastern U.S. Participants were enrolled between 2002 and 2009 from Alabama, Arkansas, Florida, Georgia, Kentucky, Louisiana, Mississippi, North Carolina, South Carolina, Tennessee, Virginia, and West Virginia. They were recruited primarily (>80%) from community health centers (CHCs), which provide basic health services to uninsured individuals, thus the cohort uniquely captures a diverse and understudied population in the Southeastern U.S. More details about the SCCS are published elsewhere. [13, 14]

In our study, we excluded SCCS participants who reported cancer at baseline (N=5,455, including 11 with MM), participants who did not self-identify as either Non-Hispanic White or Non-Hispanic Black (N=3,761), and participants who did not complete the baseline survey (N=997). We refer to Non-Hispanic White participants as White individuals and Non-Hispanic Black participants as Black individuals for brevity. Written informed consent was obtained from all SCCS participants. Vanderbilt University Medical Center and Meharry Medical College institutional review boards (IRB#010345) approved all study procedures. All research activities performed for this study adhered to the principles stated in the Declaration of Helsinki.

### 2.2. Outcomes variables

The main outcomes of interest in the current study were MM incidence and among participants with MM, all-cause mortality. Incident MM cases, defined as International Classification of Diseases (ICD-O-3 morphology 9731/3, 9732/3, and 9734/3), were identified via linkages with state-level cancer registries. We also linked data with the National Death Index (NDI) to determine all-cause mortality and vital status of all participants. Follow-up for incident cases and vital status was conducted from SCCS enrollment (between 2002-2009) until the end of follow-up; follow-up for both incident case ascertainment and for mortality varied by state between the dates of 12/31/2016 and 12/31/2019, depending on the date reported by each state registry. Participants located outside of the 12 SCCS states ended follow-up at 12/31/2019. We calculated the survival time in months, from the age at diagnosis to the age at death or the end of follow-up. We excluded four participants with MM who had less than one month of follow-up, leaving a final sample size of 162 participants with incident MM.

### 2.3. Exposure variables and covariates

SDoH factors were measured based on residence at SCCS enrollment and included the following: residential racial segregation, neighborhood deprivation, rurality, poverty, and population density. Two measures of residential racial segregation were included: the Index of Concentration at the Extremes (ICE) and the Local Exposure and Isolation (LEx/Is) index. [9] The ICE is a measure of the difference in the number of Black residents compared to White residents in an area compared to the total population. A lower score indicates an area with more White individuals or higher White privilege. The LEx/Is measures the probability that two individuals of different racial groups (Black, White) living in the same local area will interact, compared to the likelihood of interaction in the larger geographic area. A higher Lex/Is score indicates a higher likelihood of interaction and less segregation.

Rurality and poverty were defined at the census tract level and county level based on codes from the U.S. Department of Agriculture’s Economic Research Service (ERS). [15, 16] Participants were categorized as living in urban vs. rural communities based on the ERS’s rural–urban commuting area (RUCA) scores, and a persistent poverty area was defined as having high poverty (≥20%) for three consecutive decades. Population density, measured by 2010 U.S. Census Bureau, is calculated based on how many people resided in a square mile of an area in the year 2010 (RRID: SCR_011587). Neighborhood deprivation was measured using a deprivation index developed by the SCCS, using participant enrollment address and 20 Census 2000 variables linked at the census tract level. The index includes socioeconomic factors related to income, housing, education, and occupation. [17–20] All SDoH variables were categorized into quartiles based on the study sample.

Demographic characteristics (age, sex, self-identified race), BMI, annual household income, and smoking status were collected via baseline survey at SCCS enrollment. BMI was categorized as normal weight (BMI<25 kg/m^2^), overweight (BMI 25-29 kg/m^2^), and obese (BMI ≥30 kg/m^2^). Annual household income was categorized into two groups: low-income (<$15,000) and not low-income (≥$15,000). Tobacco smoking status was categorized as never smoked, formerly smoked, or currently smoking. MM cancer stage and cancer treatment data were not available for the current study.

### 2.4. Statistical analysis

Descriptive statistics were computed for demographic characteristics, smoking, and SDoH variables, including medians and interquartile ranges for continuous variables and percentages for categorical variables. Multivariable cox proportional hazard models were computed to evaluate associations between SDoH factors and MM incidence and survival. Hazard ratios and 95% confidence intervals were produced for each exposure variable for incidence and mortality with adjustment for a priori selected covariates.

Incidence rates were adjusted for age based on the SCCS cohort and were calculated as the number of MM cases divided by the number of months of follow-up (from SCCS enrollment to date of MM diagnosis). Two incidence models were computed, the first adjusting for age, race, sex, enrollment source, and household income, and the second model additionally adjusting for BMI. Among participants with MM, two mortality models were computed, the first adjusting for age, sex, race, enrollment source, and household income, and the second model additionally adjusting for smoking status and BMI.

Because other research has shown that smoking history does not increase the risk of developing MM, but is associated with mortality, we included smoking status as a covariate in the survival but not in the incidence multivariable models. [21] We evaluated potential interaction effects of race (Black and White) and BMI (“obese” (BMI ≥30) and “not obese” (BMI<30 kg/m^2^)) on the associations between the SDoH factors and cancer risk. When a significant race-exposure or BMI-exposure interaction was identified, and sample size was sufficient, further analyses stratified by race or BMI were conducted. However, because of the low number of MM cases, survival models stratified by obesity and race were not reliable estimates and not reported.

### 2.5. Data availability

The current study utilized data from the SCCS, which can be obtained by online request (https://ors.southerncommunitystudy.org/).

## 3. Results

### 3.1. Participant characteristics

Among 74,294 SCCS participants included in the study, the median age was 50 years (IQR: 46-60 years), and the majority were female (58.4%) and self-identified as Black individuals (69.8%; Table 1). Most participants (56.3%) had an annual household income of <$15,000, and about 42% were obese. About 41.9% were current smokers, 22.2% were former smokers, and 35.9% reported never smoking. About 45% of participants lived in an area with persistent poverty; most participants lived in urban areas (78.5%), and 53.8% lived in high deprivation neighborhoods.

**Table 1.** Participant Demographic Characteristics and SDoH Factors overall and among Participants with Multiple Myeloma.

|  | **Overall**<br>**Sample**<br>**N=74,294**<br>**(%)** | **Multiple Myeloma**<br>**Cases**<br>**N=162**<br>**(%)** |
| --- | --- | --- |
| **Age, Years, Median (IQR)** | 50 (46,60) | 64.0 (59,71) |
| **Sex** |  |  |
| Female | 58.4 | 63.0 |
| Male | 41.6 | 37.0 |
| **Race** |  |  |
| Black | 69.8 | 82.1 |
| White | 30.2 | 17.9 |
| **Smoking Status** |  |  |
| Never | 35.9 | 43.2 |
| Former | 22.2 | 30.2 |
| Current | 41.9 | 26.5 |
| **Income** |  |  |
| <\$15,000 | 56.3 | 58.0 |
| ≥\$15,000 | 43.7 | 42.0 |
| **BMI** |  |  |
| <25 (normal BMI) | 25.4 | 13.0 |
| 25-30 (overweight) | 30.7 | 29.6 |
| ≥30 (obese) | 42.2 | 57.4 |
| **Residential Racial Segregation (Lex/Is)** |  |  |
| Q1 (>segregation) | 21.3 | 16.7 |
| Q2 | 21.0 | 23.5 |
| Q3 | 21.9 | 25.3 |
| Q4 (<segregation)<sup>a</sup> | 21.9 | 17.9 |
| Missing | 13.9 | 16.7 |
| **Residential Racial Segregation (ICE)** |  |  |
| Q1 (>White Individuals/ >White privilege) | 22.9 | 16.0 |
| Q2 | 23.1 | 21.6 |
| Q3 | 22.8 | 24.1 |
| Q4 (<White Individuals/ <White privilege) | 23.3 | 27.8 |
| Missing | 7.9 | 10.5 |
| **Population Density** |  |  |
| Q1 (<dense) | 25.0 | 31.5 |
| Q2 | 24.9 | 29.6 |
| Q3 | 25.2 | 20.4 |
| Q4 (>dense) | 24.9 | 18.5 |
| **Persist poverty** |  |  |
| Not persistently poor area | 54.6 | 50.6 |
| Persistently poor area | 45.4 | 49.4 |
| **Deprivation Index** |  |  |
| Q1 (<deprivation) | 10.2 | 4.3 |
| Q2 | 15.4 | 15.4 |
| Q3 | 20.6 | 23.5 |
| Q4 (>deprivation) | 53.8 | 56.8 |
| **Rurality** |  |  |
| Rural | 21.5 | 27.8 |
| Urban | 78.5 | 72.2 |
| <sup>a</sup> A higher score indicates a greater likelihood of interaction of individuals of different racial groups and less segregation |  |  |

### 3.2. MM incidence

There were 162 incident MM cases (Table 1). The median age at MM diagnosis was 64 years (IQR: 59-71 years); 133 self-identified as Black individuals (82.1%) and 29 self-identified as White individuals (17.9%). Most MM cases were female (N=102; 63%), obese (57.4%) and were either current or former smokers (56.7%). In the final multivariable models, increasing neighborhood deprivation was associated with increased MM risk (Table 2). After adjustment for sociodemographic characteristics and BMI, participants living in the most deprived neighborhoods had 2.24-fold increased risk of MM compared to participants living in the least deprived neighborhoods (95% CI: 1.01-4.95). All other SDoH factors were not statistically associated with MM incidence.

**Table 2:** Multivariable Hazard Ratios (HR) and 95% Confidence Intervals (CI) for SDoH Factors and Age-adjusted Multiple Myeloma (MM) Incidence Rates (per 100,000)^a^.

|  | Incident<br>MM <sup>a</sup> | Model 1<sup>b</sup> | Model 2<sup>c</sup> |
| --- | --- | --- | --- |
|  |  | HR (95% CI) | HR (95% CI) |
| **Residential Racial Segregation (Lex/Is)** |  |  |  |
| Q1 (>segregation) | 14.1 | REF | REF |
| Q2 | 22.1 | 1.60(0.99-2.59) | 1.58(0.98-2.56) |
| Q3 | 22.5 | 1.58(0.99-2.54) | 1.54(0.96-2.48) |
| Q4 (<segregation)<sup>d</sup> | 15.8 | 1.20(0.72-2.00) | 1.18(0.70-1.96) |
| **Residential Racial Segregation (ICE)** |  |  |  |
| Q1 (>White Individuals/ >White privilege) | 13.3 | REF | REF |
| Q2 | 18.6 | 1.14(0.67-1.94) | 1.12(0.66-1.91) |
| Q3 | 20.2 | 1.11(0.64-1.93) | 1.10(0.63-1.91) |
| Q4 (<White individuals/<White privilege) | 22.1 | 1.23(0.71-2.14) | 1.24(0.71-2.16) |
| **Deprivation Index** |  |  |  |
| Q1 (<deprivation) | 7.4 | REF | REF |
| Q2 | 19.3 | **2.67(1.16-6.14)** | **2.58(1.12-5.93)** |
| Q3 | 22.3 | **2.71(1.20-6.08)** | **2.59(1.15-5.82)** |
| Q4 (>deprivation) | 20.2 | **2.29(1.04-5.08)** | **2.24(1.01-4.95)** |
| **Population Density** |  |  |  |
| Q1 (<dense) | 23.6 | REF | REF |
| Q2 | 22.3 | 0.99(0.67-1.46) | 1.01(0.68-1.49) |
| Q3 | 16.4 | 0.69(0.45-1.05) | 0.72(0.47-1.09) |
| Q4 (>dense) | 14.8 | 0.66(0.43-1.02) | 0.70(0.45-1.08) |
| **Rurality** |  |  |  |
| Rural | 24.7 | REF | REF |
| Urban | 17.7 | 0.79(0.57-1.11) | 0.81(0.58-1.14) |
| **Persistent Poverty Area** |  |  |  |
| Not persistently poor area | 17.9 | REF | REF |
| Persistently poor area | 20.8 | 1.02(0.75-1.40) | 1.03(0.75-1.41) |

<sup>a</sup> Incidence rates were adjusted for age and were calculated as the number of MM cases divided by the number of months of follow-up (from SCCS enrollment to date of MM diagnosis).
<sup>b</sup> Multivariable model 1 is adjusted for age, race, sex, enrollment source, household income. Bold indicates statistical significance at alpha level $p \leq 0.05$ .
<sup>c</sup> Multivariable model 2 is adjusted for age, race, sex, enrollment source, household income, BMI. Bold indicates statistical significance at alpha level $p \leq 0.05$ .
<sup>d</sup> A higher score indicates a greater likelihood of interaction of individuals of different racial groups and less segregation

Because obesity is a known risk factor for MM and could be associated with SDoH factors, we conducted stratified analyses for participants who were obese and those who were not obese based on a significant interaction effect by obesity and several SDoH factors. Among participants who were not obese, population density and rurality were significantly associated with MM incidence (Table 3). Specifically, participants that lived in the most densely populated areas had a reduced risk of MM compared to those who lived in the least densely populated areas (HR:0.31; 95% CI: 0.14-0.67). Additionally, those who lived in urban areas had a reduced risk compared to those who lived in rural areas (HR: 0.52; 95% CI: 0.32-0.85). Interestingly, SDoH factors were not associated with MM incidence among participants who were obese. Stratified analyses by race are included in Supplemental Table 1. Because of the small sample size among White individuals, we only report overall associations here.

**Table 3.** Multivariable Hazard Ratios (HR) and 95% Confidence Intervals (CI) for SDoH Factors and Multiple Myeloma (MM) by Obesity.

|  | **Not Obese**<br>**(BMI<30)** | **Obese**<br>**(BMI≥30)** | **P for**<br>**interaction<sup>b</sup>** |
| --- | --- | --- | --- |
|  | **HR (95% CI)<sup>a</sup>** | **HR (95% CI)<sup>a</sup>** |  |
| **Residential Racial Segregation (Lex/Is)** |  |  | **P=0.74** |
| Q1 (>segregation) | REF | REF |  |
| Q2 | 1.58(0.75-3.31) | 1.58(0.84-2.97) |  |
| Q3 | 1.89(0.93-3.85) | 1.30(0.69-2.46) |  |
| Q4 (<segregation)<sup>c</sup> | 1.47(0.69-3.14) | 0.98(0.49-1.97) |  |
| **Residential Racial Segregation (ICE)** |  |  | **P=0.37** |
| Q1 (>White individuals/ >White privilege) | REF | REF |  |
| Q2 | 1.07(0.50-2.31) | 1.20(0.58-2.49) |  |
| Q3 | 1.07(0.49-2.34) | 1.14(0.53-2.48) |  |
| Q4 (<White individuals/ <White privilege) | 0.79(0.35-1.80) | 1.77(0.83-3.77) |  |
| **Deprivation Index** |  |  | **P=0.82** |
| Q1 (<deprivation) | REF | REF |  |
| Q2 | 3.43(0.97-12.09) | 2.05(0.67-6.24) |  |
| Q3 | 3.28(0.95-11.31) | 2.17(0.75-6.31) |  |
| Q4 (>deprivation) | 2.44(0.72- 8.25) | 2.10(0.74-5.95) |  |
| **Population Density** |  |  | **P=0.03** |
| Q1 (<dense) | REF | REF |  |
| Q2 | 0.94(0.53-1.67) | 1.03(0.61-1.76) |  |
| Q3 | 0.55(0.29-1.04) | 0.84(0.48-1.48) |  |
| Q4 (>dense) | **0.31(0.14-0.67)** | 1.15(0.67-1.98) |  |
| **Rurality** |  |  | **P=0.02** |
| Rural | REF | REF |  |
| Urban | **0.52(0.32-0.85)** | 1.15(0.72-1.84) |  |
| **Persistent Poverty Area** |  |  | **P=0.80** |
| Not persistently poor area | REF | REF |  |
| Persistently poor area | 1.10(0.68-1.80) | 0.99(0.66-1.49) |  |

<sup>a</sup> Multivariable model is adjusted for age, race, sex, enrollment source, household income. Bold indicates statistical significance at alpha level $p \leq 0.05$ .
<sup>b</sup> P for interaction between obesity and each SDoH factor was calculated in the main model (Table 2). It indicates the homogeneity or heterogeneity of the exposure (e.g., population density) effect across the strata (e.g., BMI<30 vs BMI≥30).
<sup>c</sup> A higher score indicates a greater likelihood of interaction of individuals of different racial groups and less segregation

### 3.3. MM survival

Among the 162 participants diagnosed with MM, 58% died during follow-up with a survival range of 1 to 153 months (Mean: 35.4 months (SD: 34.3 months)). In the final multivariable model, residential racial segregation (Lex/Is) was associated with mortality (Table 4). After adjustment for sociodemographic characteristics and BMI, living in the least segregated areas was associated with 2.21-fold increased mortality compared to living in the most segregated areas (95% CI: 1.03-4.74). Based on this finding, we tested for an interaction between residential racial segregation (Lex/Is) and income and found that it was not statistically significant (p=0.18). All other SDoH factors were not statistically associated with mortality.

**Table 4.** Multivariable Hazard Ratios (HR) and 95% Confidence Intervals (CI) for SDoH Factors and Survival among Participants with Multiple Myeloma (N=162)

|  | Deaths<sup>a</sup> | Model 1<sup>b</sup> | Model 2<sup>c</sup> |
| --- | --- | --- | --- |
|  |  | HR (95% CI) | HR (95% CI) |
| **Residential Racial Segregation (Lex/Is)** |  |  |  |
| Q1 (>segregation) | 12 | REF | REF |
| Q2 | 20 | 1.42(0.69-2.92) | 1.84(0.87-3.90) |
| Q3 | 28 | 1.55(0.78-3.10) | 1.88(0.93-3.78) |
| Q4 (<segregation)<sup>d</sup> | 17 | 1.87(0.88-3.97) | **2.21(1.03-4.74)** |
| **Residential Racial Segregation (ICE)** |  |  |  |
| Q1 (>White individuals/ >White privilege) | 13 | REF | REF |
| Q2 | 22 | 1.21(0.56-2.64) | 1.23(0.57-2.68) |
| Q3 | 26 | 1.23(0.56-2.71) | 1.14(0.51-2.54) |
| Q4 (<White individuals/ <White privilege) | 24 | 0.87(0.38-1.96) | 0.72(0.31-1.67) |
| **Deprivation Index** |  |  |  |
| Q1 (<deprivation) | -<sup>e</sup> | REF | REF |
| Q2 | -<sup>e</sup> | 1.69(0.47-6.06) | 1.31(0.36-4.81) |
| Q3 | 24 | 2.71(0.79-9.27) | 2.05(0.59-7.12) |
| Q4 (>deprivation) | 54 | 2.12(0.64-7.06) | 1.53(0.44-5.29) |
| **Population Density** |  |  |  |
| Q1 (<dense) | 33 | REF | REF |
| Q2 | 26 | 0.84(0.49-1.42) | 0.75(0.43-1.30) |
| Q3 | 16 | 0.63(0.34-1.15) | 0.53(0.29-1.00) |
| Q4 (>dense) | 19 | 1.18(0.64-2.19) | 0.98(0.51-1.86) |
| **Rurality** |  |  |  |
| Rural | 30 | REF | REF |
| Urban | 64 | 0.82(0.53-1.27) | 0.75(0.47-1.19) |
| **Persistent Poverty Area** |  |  |  |
| Not persistently poor area | 48 | REF | REF |
| Persistently poor area | 46 | 0.99(0.66-1.51) | 1.00(0.65-1.52) |
| <sup>a</sup> All-cause mortality reported as number of deaths. |  |  |  |
| <sup>b</sup> Adjusted for age, sex, race, enrollment source, household income. Bold indicates statistical significance at alpha level p≤0.05. |  |  |  |
| <sup>c</sup> Adjusted for age, sex, race, enrollment source, household income, smoking, and BMI. Bold indicates statistical significance at alpha level p≤0.05. |  |  |  |
| <sup>d</sup> A higher score indicates a greater likelihood of interaction of individuals of different racial groups and less segregation |  |  |  |
| <sup>e</sup> Cell count too small to report |  |  |  |

## 4. Discussion

To our knowledge, we are the first to show a significant association between residing in neighborhoods with high deprivation based on geocoding and MM risk among a large cohort of Black and White, primarily low-income, individuals in the Southeastern U.S. Living in areas with high population density was associated with reduced MM incidence among those with lower BMI (<30 kg/m^2^) indicating that built environment of access to healthy foods and walkability may decrease MM risk. Walkability and access to healthy foods have been shown to be associated with reduced obesity in urban settings; however, associations vary by socioeconomic status and other social factors such as crime rates. [22–24] Neighborhood walkability has also been shown to be inversely associated with BMI among cancer survivors. [25] These findings emphasize the need for reducing health disparities in cancer burden faced in underserved communities.

Although there are limited data examining SDoH and environmental exposures associated with MM incidence, in another study examining county-level geospatial distribution of MM incidence in the U.S., Cheung et al. discovered nonrandom distribution of MM cases, with specific clusters of incidence rates in different regions in the U.S. [22] They also found significant associations with race (Black vs. White individuals) and MM incidence clustering, suggesting that racial distribution may explain some of the distinct MM incidence clustering they observed. The findings from that study and our study suggest that MM risk is complex and includes a multitude of individual and social factors.

Based on SEER data from 2015-2021, the five-year relative survival rate of MM in the U.S. is approximately 62%. [1] According to SEER data, Black individuals are more likely to die from MM compared to White individuals with MM. Among participants in our study who developed MM, 58% died during follow-up and there were no significant differences in mortality by race. As we know from previous research, staging and cytogenetic factors play a role in MM survival; however, we were limited in the current analyses to account for these factors.

The literature examining SDoH and mortality among people with MM is very limited. One study by Hong and colleagues, utilizing SEER data from 2006-2016 suggests that income and neighborhood deprivation may have interactive effects on mortality. [23] Among approximately 7,000 SEER participants with MM, they found that low income was associated with increased all-cause mortality compared to not having low income, but that this relationship varied across levels of neighborhood deprivation. Specifically, they found that among people residing in the highest deprivation areas, having low income was not associated with increased mortality, as it was in all other groups. In our study, we found that neighborhood deprivation was not statistically associated with mortality risk. These findings suggest that the interplay between individual low-income status and indices of neighborhood deprivation on mortality risk among people with MM is complex and warrants further investigation.

SEER data shows that MM incidence rates are substantially higher among Non-Hispanic Black individuals compared to White individuals. [1] And in our study, most of the participants who developed MM self-identified as Black individuals. We computed separate models by race (Supplemental Table 1) but did not report the results in the main findings due to unstable estimates given the low sample size by race. We did, however, attempt to account for racial differences by adjustment in our multivariable models. As there is a genetic component of MM risk in both Black and White individuals [24], one limitation of the study was that we did not have ancestral or genetic information on study participants, and self-reported race was used in the analyses instead. Interestingly, none of the incident MM cases in our study reported a family history of MM, which has been shown to be associated with a 2-fold increased risk. [25] Additionally, there were only two incident cases who did not self-identify as Black or White; therefore, we did not include individuals outside of those two racial groups in these analyses.

Because obesity is a well-established risk factor for MM and is associated with its precursor, MGUS, in the current study, we considered obesity and examined MM incidence separately for those who were and were not obese. In these stratified analyses, we discovered that living in areas with higher population density and living in urban areas were associated with decreased MM risk among participants who were not obese, but these associations were not seen among people who were obese. A study led by Landgren and colleagues conducted among a smaller (N=1,996) randomly selected sample from the SCCS cohort found a 1.8-fold increased risk of MGUS among participants who were obese compared to participants who were not obese, independent of race. [26] These findings suggest that investigating MM risk should involve a multifactorial approach that includes genetic factors, lifestyle factors, and SDoH.

There were a few additional limitations in the current study. First, we did not account for MM staging or MM treatment in our analyses, as this information was not available from the state-level cancer registries or NDI. We acknowledge that treatment differences and delays in treatment have been shown to worsen outcomes, and to perpetuate racial disparities. [27] Second, SDoH geographic exposures in the current study were linked to participant address at the time of SCCS enrollment, and do not account for geographic movement of participants over time. Third, exposure variables based on baseline survey data are subject to recall bias. However, linkage with the state-level cancer registries and the NDI provided a vigorous approach to ascertaining our primary outcomes of incident MM and deaths.

## 5. Conclusion

In summary, in the current study, we examined SDoH factors associated with MM incidence and survival among a large cohort of predominantly low-income Black and White individuals residing in the Southeastern U.S. We discovered that higher neighborhood deprivation was associated with increased MM risk. Additionally, we found that residential racial segregation was significantly associated with mortality among MM cases. These findings suggest that SDoH factors may significantly impact MM risk and survival among low-income populations in the Southern U.S. and should be considered when developing strategies to reduce MM burden and disparities among populations with lower socioeconomic backgrounds.

**Supplemental Table 1.** Multivariable Hazard Ratios (HR) and 95% Confidence Intervals (CI) for Multiple Myeloma (MM) by SDoH Factors and Race.

|  | White | Black |  |
| --- | --- | --- | --- |
|  | HR (95% CI)<sup>a</sup> | HR (95% CI)<sup>a</sup> | P-value for interaction<sup>b</sup> |
| **Residential Racial Segregation (Lex/Is)** |  |  |  |
| Q1 (>segregation) | REF | REF |  |
| Q2 | 1.56(0.39-6.26) | 1.59(0.96-2.65) |  |
| Q3 | 1.72(0.45-6.68) | 1.52(0.92-2.52) |  |
| Q4 (<segregation)<sup>c</sup> | 1.65(0.44-6.25) | 1.07(0.61-1.89) | P=0.85 |
| **Residential Racial Segregation (ICE)** |  |  |  |
| Q1 (>White individuals/ >White privilege) | REF | REF |  |
| Q2 | 1.36(0.57- 3.26) | 0.95(0.50-1.82) |  |
| Q3 | 0.98(0.22- 4.31) | 0.97(0.52-1.82) |  |
| Q4 (<White individuals/ <White privilege) | 3.37(0.76-14.98) | 1.06(0.57-1.97) | P=0.53 |
| **Deprivation Index** |  |  |  |
| Q1 (<deprivation) | REF | REF |  |
| Q2 | 1.55(0.38- 6.27) | **3.07(1.05-8.95)** |  |
| Q3 | 2.47(0.66- 9.17) | 2.52(0.89-7.15) |  |
| Q4 (>deprivation) | 3.80(1.02-14.13) | 2.06(0.75-5.64) | P=0.10 |
| **Population Density** |  |  |  |
| Q1 (<dense) | REF | REF |  |
| Q2 | 1.02(0.43-2.40) | 1.00(0.65-1.54) |  |
| Q3 | 0.95(0.33-2.74) | 0.68(0.43-1.07) |  |
| Q4 (>dense) | 0.87(0.27-2.74) | 0.67(0.42-1.07) | P=0.90 |
| **Rurality** |  |  |  |
| Rural | REF | REF |  |
| Urban | 0.51(0.24-1.08) | 0.90(0.62-1.30) | P=0.23 |
| **Persistent Poverty Area** |  |  |  |
| Not persistently poor area | REF | REF |  |
| Persistently poor area | **2.17(1.00-4.71)** | 0.92(0.66-1.28) | **P=0.03** |

<sup>a</sup> Multivariable model is adjusted for age, sex, enrollment source, household income and BMI.
Bold indicates statistical significance at alpha level $p \leq 0.05$ .
<sup>b</sup> P for interaction between obesity and each SDoH factor was calculated in the main model (Table 2). It indicates the homogeneity or heterogeneity of the exposure (e.g., population density) effect across the strata (e.g., white vs black)
<sup>c</sup> A higher score indicates a greater likelihood of interaction of individuals of different racial groups and less segregation

## CRediT Roles

Anna Junkins: Conceptualization; Investigation; Methodology; Writing-original draft; Writing-review & editing; Wanqing Wen: Conceptualization; Formal analysis; Investigation; Writing-review & editing, Loren Lipworth: Conceptualization; Methodology; Writing-original draft; Writing-review & editing, Xijing Han: Data curation; Formal analysis; Writing-review & editing, Heather Munro: Data curation; Formal analysis; Writing-review & editing, Michael T. Mumma: Data curation; Formal analysis; Writing-review & editing, Martha J. Shrubsole: Data curation; Funding acquisition, Methodology; Project Administration; Writing-review & editing, Wei Zheng: Conceptualization; Data curation; Project Administration; Writing-review & editing; Eden Biltibo: Methodology; Writing-review & editing; and Staci Sudenga: Conceptualization; Data curation; Formal analysis; Methodology; Supervision; Writing-original draft; Writing-review & editing.

## Data Availability

The data that support the findings of this study are available by the Southern Community Cohort Study. Restrictions apply to the availability of these data, which were used under license for this study. Data are available at (https://ors.southerncommunitystudy.org/ with the permission of the Southern Community Cohort Study.

